# Stochastic discrete epidemic modeling of COVID-19 transmission in the Province of Shaanxi incorporating public health intervention and case importation

**DOI:** 10.1101/2020.02.25.20027615

**Authors:** Sanyi Tang, Biao Tang, Nicola Luigi Bragazzi, Fan Xia, Tangjuan Li, Sha He, Pengyu Ren, Xia Wang, Zhihang Peng, Yanni Xiao, Jianhong Wu

## Abstract

Before the lock-down of Wuhan/Hubei/China, on January 23^rd^ 2020, a large number of individuals infected by COVID-19 moved from the epicenter Wuhan and the Hubei province due to the Spring Festival, resulting in an epidemic in the other provinces including the Shaanxi province. The epidemic scale in Shaanxi was comparatively small and with half of cases being imported from the epicenter. Based on the complete epidemic data including the symptom onset time and transmission chains, we calculate the control reproduction number (1.48-1.69) in Xi’an. We could also compute the time transition, for each imported or local case, from the latent, to infected, to hospitalized compartment, as well as the effective reproduction number. This calculation enables us to revise our early deterministic transmission model to a stochastic discrete epidemic model with case importation and parameterize it. Our model-based analyses reveal that the newly generated infections decay to zero quickly; the cumulative number of case-driven quarantined individuals via contact tracing stabilize at a manageable level, indicating that the intervention strategies implemented in the Shaanxi province have been effective. Risk analyses, important for the consideration of “resumption of work”, show that a large second outbreak is expected if the level of case importation remains at the same level as between January 10^th^ and February 4^th^ 2020. However, if the case importation decreases by 30%, 60% and 90%, the second outbreak if happening will be of small-scale assuming contact tracing and quarantine/isolation remain as effective as before. Finally, we consider the effects of intermittent inflow with a Poisson distribution on the likelihood of multiple outbreaks. We believe the developed methodology and stochastic model provide an important model framework for the evaluation of revising travel restriction rules in the consideration of resuming social-economic activities while managing the disease control with potential case importation.

## Introduction

Coronaviruses are single-stranded RNA viruses, with a spherical shape and crown-like spines, from which their name derives. They generally cause mild respiratory infections but can sometimes be fatal. Since their first discovery and molecular characterization, three major outbreaks have occurred, in mainland China in 2003, in the Kingdom of Saudi Arabia in 2012 and in South Korea in 2015. The fourth large-scale outbreak, caused by a novel coronavirus termed as COVID-19 and with the capital city Wuhan of the Hubei province in China as the epicenter, has spread out globally [2, 6, 8, 11], causing approximately 80,000 cases and 2,400 deaths as of February 22^nd^ 2020 [13].

Since January 23^rd^ 2020, the Chinese authorities have been implementing unprecedented and increasingly stringent public health interventions, including a complete lock-down of Wuhan and neighboring cities, strict contact tracing and travel restrictions in the epicenter and across the country [13]. Incorporating these interventions into the classical epidemic models has resulted in reliable estimates of epidemiological characteristics of COVID-19 transmission in the epicenter and accurate near-casting the epidemic trend and peak time [18-20], in comparison with other earlier modeling studies focusing on analyzing the transmission risk based on data at the early outbreak stage [1, 3, 4, 17]. One important lesson gained from our recent experience suggests the need of updating the model parameters and model setting to reflect the rapid improvement of screening, diagnosing and testing techniques/procedures, accompanied by intensive contact tracing, quarantine and isolation.

The feasibility of enhancing model capacity to assess transmission risk and evaluating intervention effectiveness has also been substantially increased due to the improvement of the quality of the data, specially from other Chinese cities and provinces where the complete lock-down of major cities such as Wuhan in the Hubei province has prevented a significant portion of infection importation, so the reduction of imported cases coupled with intensive contact tracing produced some high quality data with rather complete information on symptom onset time and transmission-chain, as will be discussed in the data section below. This enhanced model capacity for near-casting is particularly important, since the gradually relaxing of the travel restriction demands proactive- and real-time assessment of the risk of a potential second outbreak in other Chinese cities. An objective of our study is to provide this assessment, and to use this assessment along with the incorporation of feasible public health interventions into our stochastic disease transmission model to suggest the scale of interventions for these cities to prevent a second outbreak. As COVID-19 spread has occurred in many regions globally, we believe our study provides an important tool to inform public health decisions regarding travel restriction and social distancing in these regions according to their public health capacity to maintain an effective contact tracing, quarantine and isolation procedure.

In order to achieve the above research objectives, we examine the transmission characteristics of the COVID-19 epidemic in other areas outside Hubei province and quantify the risk of case importation into the current outbreak which is expected to end soon, and into a potential secondary outbreak due to the “resumption of work” in these areas. We focus on the effectiveness and implementation timeliness of prevention and control strategies, as well as the choice of timing for the “resumption of work” and its impact on the risk of a secondary outbreak.

Our analysis is based on the complete “tracking and recording” information of imported cases and local cases detected in the province of Shaanxi province [7], as this allows us to generate a data set that includes the time series of daily latent, infected and hospitalized cases. Since the outbreak scale in the Province of Shanxi was relatively small and this is subject to stochasticity in both case importation and local cases generated by the importation and since the Provincial data has detailed information on symptom onset and serial intervals, we adopt a method developed by Nishiura and Chowell [15, 16] to estimate the effective reproduction number, and we revise some of our early deterministic models to a novel discrete stochastic model to address the key issues relevant to preventing a secondary outbreak of COVID-19 in other provinces except for Hubei, due to the anticipated gradually relaxing of travel restriction and returning to normal social and economic activities.

## Methods

### Data

We obtained the data of laboratory-confirmed 2019-nCov cases in Shaanxi province from the “National Health Commission” of the People’s Republic of China and the “Health Commission” of the Shaanxi Province [7, 13]. It is worth mentioning that very detailed data information for each confirmed case has been released. This strict contact tracing strategy gave detailed information of illness onset, the first medical visit, quarantined period and confirmation. Then the data do not only include numbers, such as the number of newly reported cases, the cumulative number of reported confirmed cases, the cumulative number of cured cases and the number of death cases, and the cumulative numbers of quarantined and suspected cases (shown in Figure 1(A)), but also include the information such as the dates of illness onset, first medical visit, isolation, laboratory confirmation and discharge, and/or date of coming into Shaanxi (for imported cases). The data were released and analyzed anonymously.

**Figure 1.**
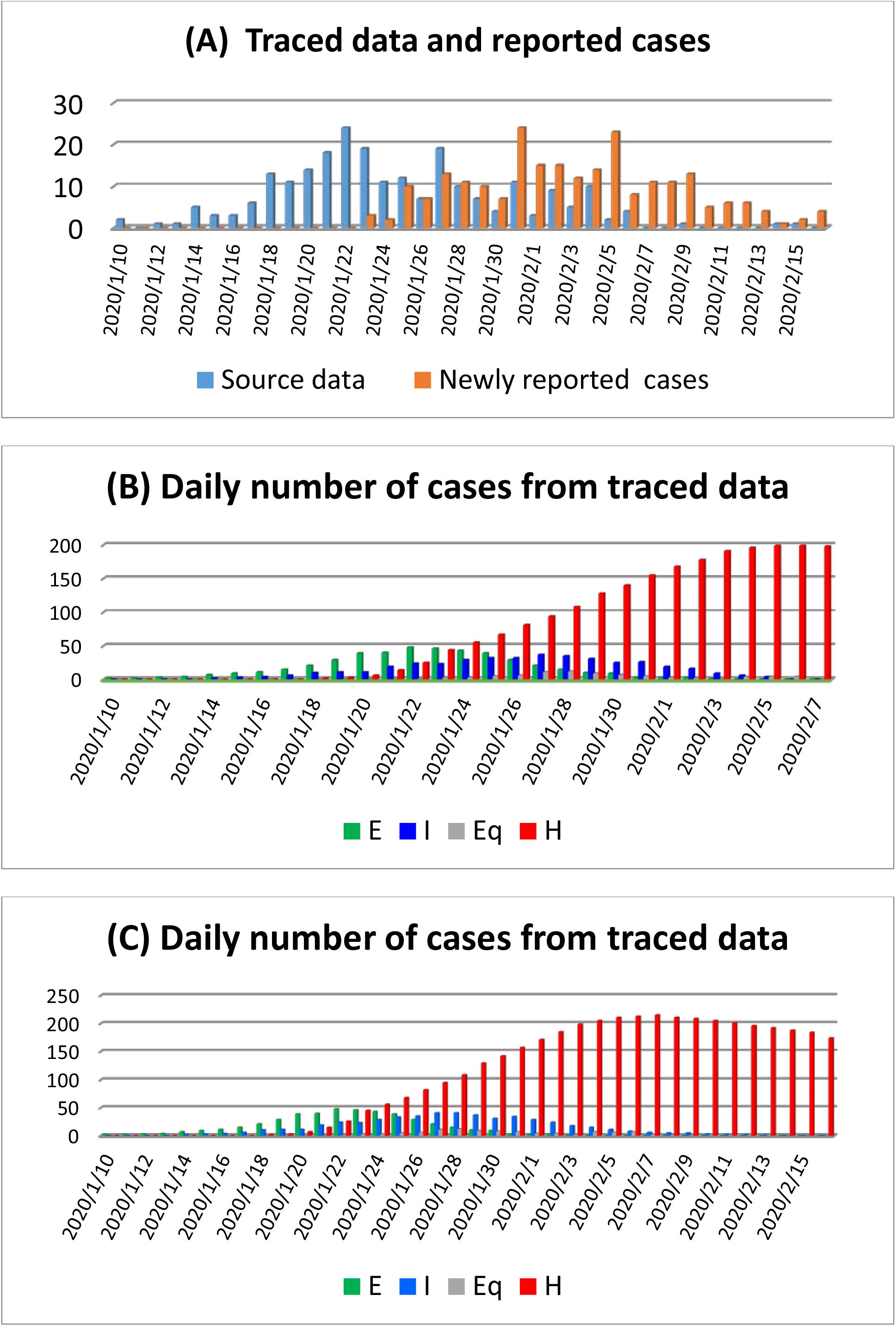
(A) Newly reported cases and daily new cases from traced data. The daily number of exposed, infected and hospitalized individuals in Shaanxi province based on the traced data by February 9^th^ (B), and by February 16^th^ 2020 (C)

Based on the recorded information, we can get the daily number of latent, infected and hospitalized individuals, and we call these data sets as traced data, as shown in Figure 1(A-C). Note that that daily number of hospitalized cases means the first medical visit and then being quarantined rather than daily number of confirmed cases. Further, we collect the data of population mobility from the website of Baidu Qianxi (http://qianxi.baidu.com). It contains the trend of inflow of population to Shaanxi before the 2020 Spring Festival and after 2019 Spring Festival, as shown in Figure 2(B).

**Figure 2.**
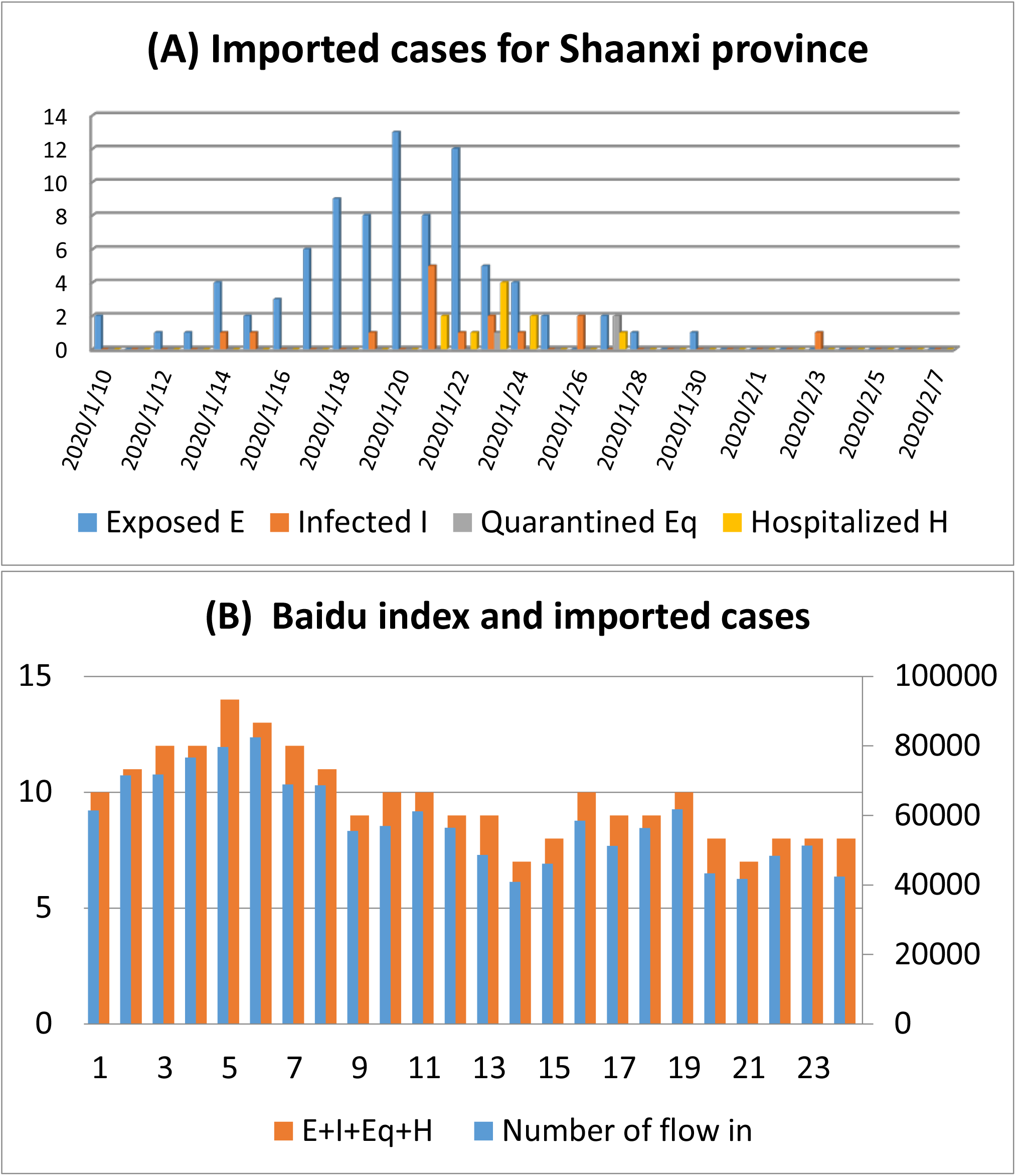
(A) The imported cases for the four compartments during Jan 10th to Feb 7th, 2020. (B) The total imported cases and number of flow into the Shaanxi province from other regions from January 10^th^ to February 3^rd^.

### Estimation of the effective reproduction number *R*(*t*)

Let *M*_*t*_ and 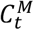 be the number of newly imported cases and the number of newly confirmed imported cases on day *t* in Shaanxi province. *T*_*M*_ represents the duration from importation to confirmation for imported cases. For *k* consecutive days j = *t*_1_, …, *t*_*k*_, *M*_*j*_ is assumed to follow the Poisson distribution with mean *λ*_*j*_ (that has to be estimated). Given the daily number of newly confirmed imported cases 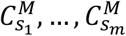 on *m* consecutive days *s*_1_, …, *s*_*m*_ and the probability *p*_*ij*_ = *P*(*i* − *j* ≤ *T*_*M*_ < *i* − *j* + 1) that an imported case entered Shaanxi province on day *j* and was confirmed on day *i*. We assume that for each *j*, 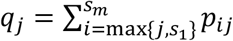 is positive. Then parameters 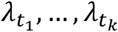 can be estimated by 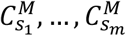 and *p*_*ij*_ through deconvolution method. We use the Richardson-Lucy iterative algorithm [5] to solve this problem. The procedure is iterative according to the following formulas:

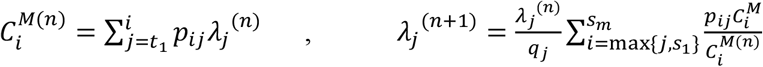

where 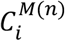 and *λ*_*j*_^(*n*)^ are fitted values of 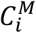 and *λ*_*j*_ in the *n*-^th^ iteration respectively. We stop the iteration when the error of fitting

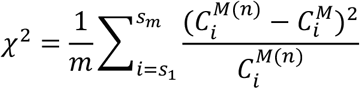

becomes small and the value of 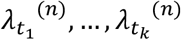 are reasonable. Because the method above requires that 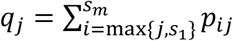 is positive, which means that at least some fraction of imported cases who entered Shaanxi province on day *j* would be confirmed during the period *s*_1_, …, *s*_*m*_, we then can determine the proper *t*_1_ and *t*_2_ such that *q*_*j*_ > 0 for *t*_1_ ≤ *j* ≤ *t*_2_ in terms of the distribution of *T*_*M*_. By using the same method, given the daily number of confirmed local cases and the probability distribution of the duration from infection to confirmation for local cases, we can also estimate the daily number of newly infected local cases *L*_*t*_ on day *t* during some time period.

To estimate the effective reproduction number *R*(*t*), we make an adaptation of the method in [15,16] to allow for imported cases. With the same notations and assumptions above, for *R*(*t*) we have

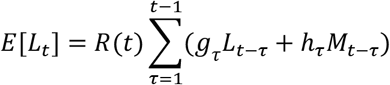

where *g*_*τ*_ = G(τ) − G(τ − 1) is the discretized distribution of the serial interval with G(τ) being the cumulative distribution function. Let *T*_*h*_ be the duration from importation of a primary imported case to infection of the secondary local case, *h*_*τ*_ = H(*τ*) − H(τ − 1) is the discretized distribution of *T*_*h*_ with H(*τ*) being the cumulative distribution function.

## The model

On the basis of the clinical progression of the disease, epidemiological status of the individuals, and intervention measures we stratify the populations as susceptible (*S*), exposed (*E*), infected (*I*), hospitalized (*H*), recovered (*R*) compartments, the quarantined susceptible (*S*_*q*_) and quarantined suspected individuals (*B*). The flow diagram is shown in Figure 3. Note that here the model framework extends the model structure in our previous study by including the quarantined suspected compartment [18-20], which consists of exposed infectious individuals resulting from contact tracing and individuals with influenza like illness and common fever needing clinical medication. The deterministic model is replaced by a discrete system with stochastic importation since the case numbers are relatively small with most of the early cases being imported in Shaanxi province [10, 12]. This modelling approach can well describe demographic stochasticity. The model equations are

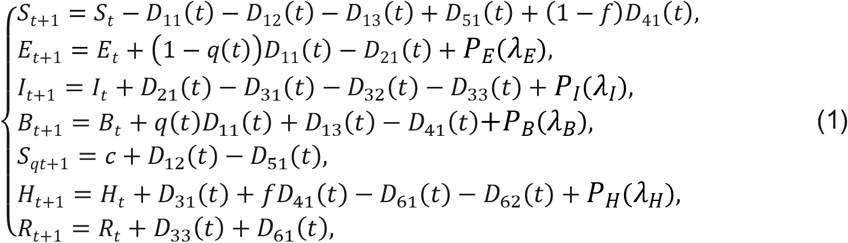

where the random variables in (1) can be defined by binomial *Bin(n, p)* distributions as follows [10,12]:

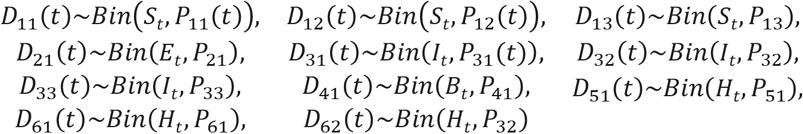

with probabilities:

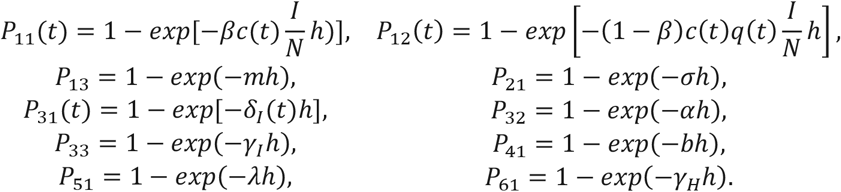

**Figure 3.**
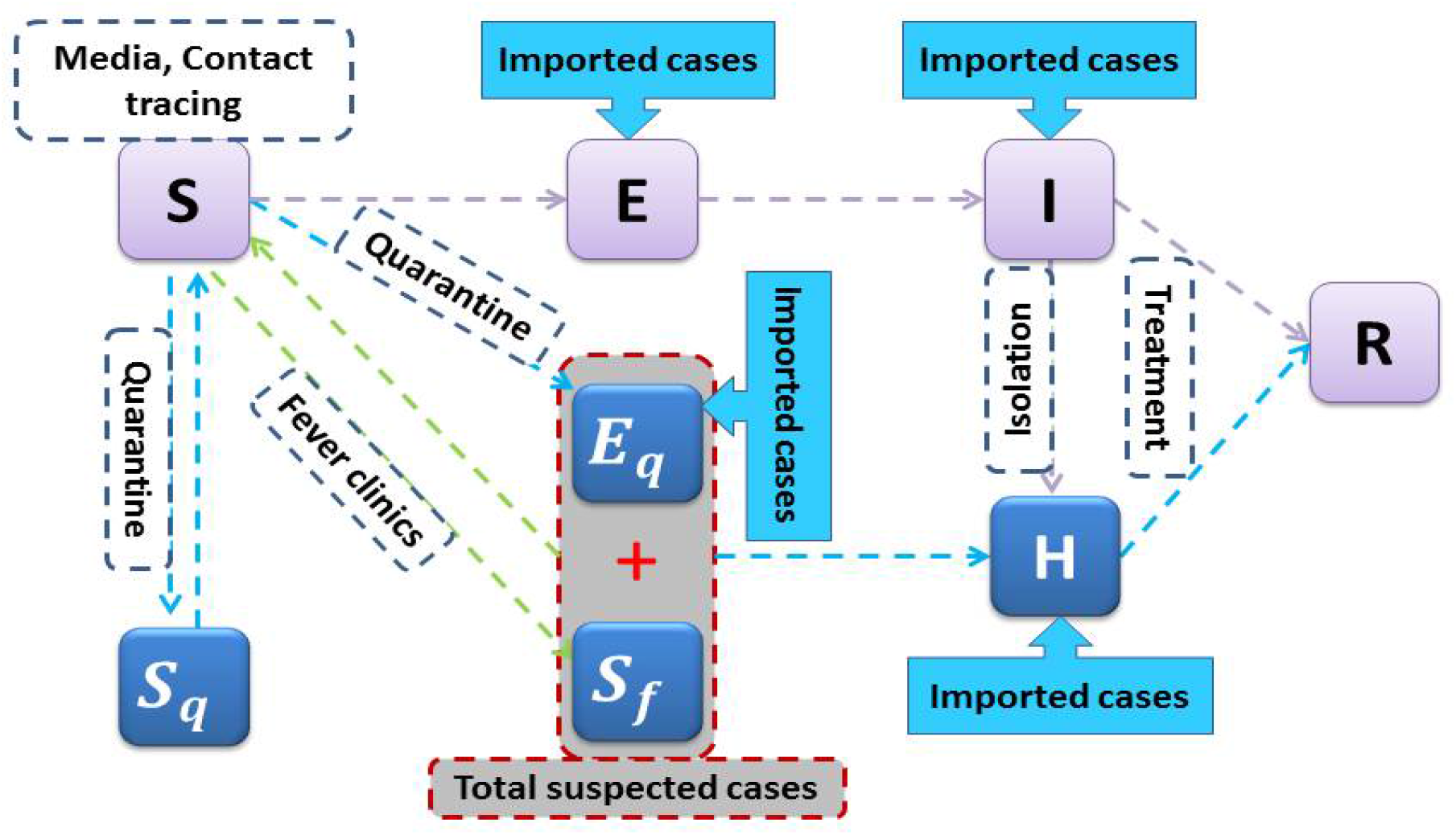
Diagram of the model adopted in the study for simulating the COVID-19 infection. Interventions including intensive contact tracing followed by quarantine and isolation are indicated. The gray compartment means suspected case compartment consisting of contact tracing Eq and fever clinics.

Let constant *m* be the transition rate from susceptible class to the suspected compartment via general clinical medication due to fever or illness-like symptoms. The suspected individuals leave this compartment at a rate of *b*, with a proportion, *f*, if has been confirmed to be infected by the COVID-19, going to the hospitalized compartment, whilst the other proportion, *1-f*, has been proven to be not infected by the COVID-19 and goes back to the susceptible class once recovery. Let the transmission probability be β and the contact rate be *c*. With contact tracing, we assume a proportion, q, of individuals exposed to the virus is quarantined, and can either move to the compartment B or S_q_ at a rate of βcq or (1 – β)cq), depending on whether they are infected or not, while the other proportion, 1 – q, consists of individuals exposed to the virus who are missed from contact tracing and move to the exposed compartment E at a rate of βc(1 − q) once infected or stay in compartment S otherwise. All definitions of variables and parameters have been listed in Table 1.

**Table 1:** Parameter estimates for the 2019-nCov epidemics in Shaanxi Province, China.

| Parameter | Definitions | Shaanxi | Source |
| --- | --- | --- | --- |
| $c_0$ | Contact rate at the initial time | 15 | 9 |
| $c_b$ | Minimum contact rate under the current control strategies | 6.0371 | Estimated |
| $r_1$ | Exponential decreasing rate of contact rate | 0.051 | Estimated |
| $\beta$ | Probability of transmission per contact | 0.21 | Estimated |
| $q_0$ | Quarantined rate of exposed individuals at the initial time | 0 | Data |
| $q_m$ | Maximum quarantined rate of exposed individuals under the current control strategies | 0.5228 | Estimated |
| $r_2$ | Exponential increasing rate of quarantined rate of exposed individuals | 0.0799 | Estimated |
| $m$ | Transition rate of susceptible individuals to the suspected class | 0.007 | Estimated |
| $b$ | Detection rate of the suspected class | 0.0397 | Estimated |
| $f$ | Confirmation ratio: Transition rate of exposed individuals in the suspected class to the quarantined infected class | 0.01 | Estimated |
| $\sigma$ | Transition rate of exposed individuals to the infected class | 1/2.38 | Data |
| $\lambda$ | Rate at which the quarantined uninfected contacts were released into the wider community | 1/12 | 9 |
| $\delta_{I0}$ | Initial transition rate of symptomatic infected individuals to the quarantined infected class | 1/4.01 | Data |
| $\delta_{If}$ | Fastest diagnose rate | 0.4076 | Estimated |
| $r_3$ | Exponential decreasing rate of diagnose rate | 0.0832 | Estimated |
| $\gamma_I$ | Recovery rate of infected individuals | 0.0497 | 9 |
| $\gamma_H$ | Recovery rate of quarantined infected individuals | 0.0199 | Estimated |
| $\alpha$ | Disease-induced death rate | 0 | Data |
| Initial values | Definitions | Shaanxi | Source |
| $S(0)$ | Initial susceptible population | $9.9895 \times 10^4$ | Estimated |
| $E(0)$ | Initial exposed population | 0 | Data |
| $I(0)$ | Initial infected population | 0 | Data |
| $B(0)$ | Initial suspected population | 0 | Data |
| $S_q(0)$ | Initial quarantined susceptible population | 0 | Data |
| $H(0)$ | Initial quarantined infected population | 0 | Data |
| $R(0)$ | Initial recovered population | 0 | Data |

The imported latent, infected and suspected and hospitalized cases for each day can be described by Poisson distributions *P*_*E*_(*λ*_*E*_), *P*_*I*_(*λ*_*I*_), *P*_*B*_(*λ*_*B*_) and *P*_*H*_(*λ*_*H*_) with parameters *λ*_*E*_, *λ*_*I*_, *λ*_*B*_ and *λ*_*H*_, respectively. Therefore,we have

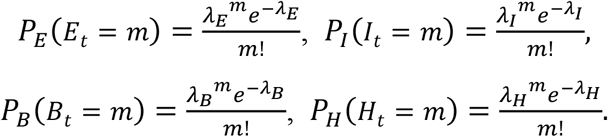

Fortunately, the number of imported cases to the above four compartments in Shaanxi province can be fully determined according to the traced data, as shown in Figures 2(A).

Since the implementation of the lock-down strategy in Wuhan on January 23^rd^ 2020, the prevention strategies in other parts of mainland China have been continuously strengthened. In order to truly describe the constantly strengthening of control strategies, we should consider the contact rate, quarantine rate, diagnosis rate and cure rate as the functions of time *t*. In this model, we assume that the contact rate c(t) is a decreasing function with respect to time t, given by

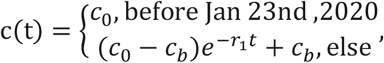

where c_0_ denotes the baseline contact rate at the initial time (i.e. before January 23^rd^, 2020) with c(0) = c_0_, c_b_ denotes the minimum contact rate under the current control strategies with lim_t→∞_ c(t) = c_b_, where c_b_ < c_0_, and r_1_ denotes the exponential decreasing rate in the contact rate.

Similarly, to characterize the enhanced contact tracing we define *q*(*t*) as an increasing function with respect to time *t*, written as

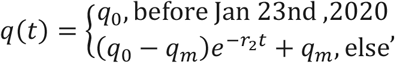

where *q*_0_ is the initial quarantined rate of exposed individuals with *q*(0) = *q*_0_ = 0 in Shaanxi province, *q*_*m*_ is the maximum quarantined rate under the current control strategies with lim_*t*→∞_ *q*(*t*) = *q*_*m*_ and *q*_*m*_ > *q*_0_, and *r*_2_ represents the exponential increasing rate in quarantined rate. We also set the transition rate *δ*_*I*_(*t*) as an increasing function with respect to time *t*, thus the detection period 1/*δ*_*I*_(*t*) is a decreasing function of *t* with the following form:

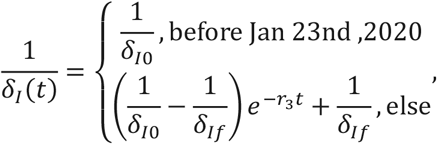

where δ_I0_ is the initial rate of confirmation, *δ*_*If*_ is the fastest confirmation rate, and *r*_3_ is the exponential decreasing rate of the detection period. Definitely, *δ*_*I*_(0) = *δ*_*I*0_ and lim_*t*→∞_ *δ*_*I*_(*t*) = *δ*_*If*_ with *δ*_*If*_ > *δ*_*I*0_. Let *T*_*c*_ be the time point at which stringent control measures are introduced on Jan 23nd, 2020. Until February 4^th^ 2020, there was no cured case in Shaanxi province, so the cure rate *γ*_*H*_ should be zero before February 4^th^ 2020. The number of discharged patients has increased rapidly since February 7^th^ 2020. Therefore, the cure rate is defined as a constant piecewise function based on the real situation.

### Simulation of the resumption of work

When simulating the resumption of work, we assume that the movement trend of Shaanxi population after resumption of work is the same as that in same period of 2019. Let the average proportions of E, I, E_q,_H that flow in the Shaanxi during January 17^th^ to 22^nd^, 2020 be p_E_, p_I_, p_B_, p_H_, which can be calculated by the data of imported cases and population mobility. Let the proportions of E, I, E_q,_H that flow in Shaanxi province after resumption of work be 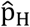. We initially assume that 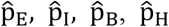 are the same as those of p_E_, p_I_, p_B_, p_H_, when the resumption of work begins on February 24th, then we consider the proportions 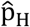 may reduce 30%, 60% and even 90% corresponding to weaker and weaker epidemic.

## Main results

### Data implications

From the detailed description about traced data, we can know when and what is the state of each infected person and obtain the number of who are in latent (or quarantined latent), infected and hospitalized classes, as shown in Figure 1, which shows the evolution process of the epidemic in Shaanxi province. Figure 1(A) gives both newly reported cases and new cases from traced data, and comparison of two time series indicates that the new cases from traced data is about 7 days ahead of the reported cases data. In particular, Figure 1(B) and (C) give the epidemic status of cases confirmed before February 7^th^ and 15^th^, respectively. Note that although Shaanxi province began to release the case numbers on January 23^rd^, 2020, the first imported latent case came to Shaanxi as early as January 10^th^, 2020, and the majority of imported cases were latent and came into Shaanxi before the end of January, 2020, as shown in Figure 2(A). Figure2(B) shows the correlation between the total imported cases and the number of population flow into the Shaanxi province, from which we calculate the proportion of those inflows that are infected.

We can also illustrate the spatial distribution of imported cases (circles) and local cases (squares) and disease evolution based on the traced data. It follows from Figure 4 that most cases are in Xi’an city, few cases are in the northern part of Shaanxi while relatively many cases are in the southern part of Shaanxi, which is adjacent to Hubei province. Therefore, we can estimate the impact of population migration on the epidemic situation in the neighboring areas of Hubei Province.

**Figure 4.**
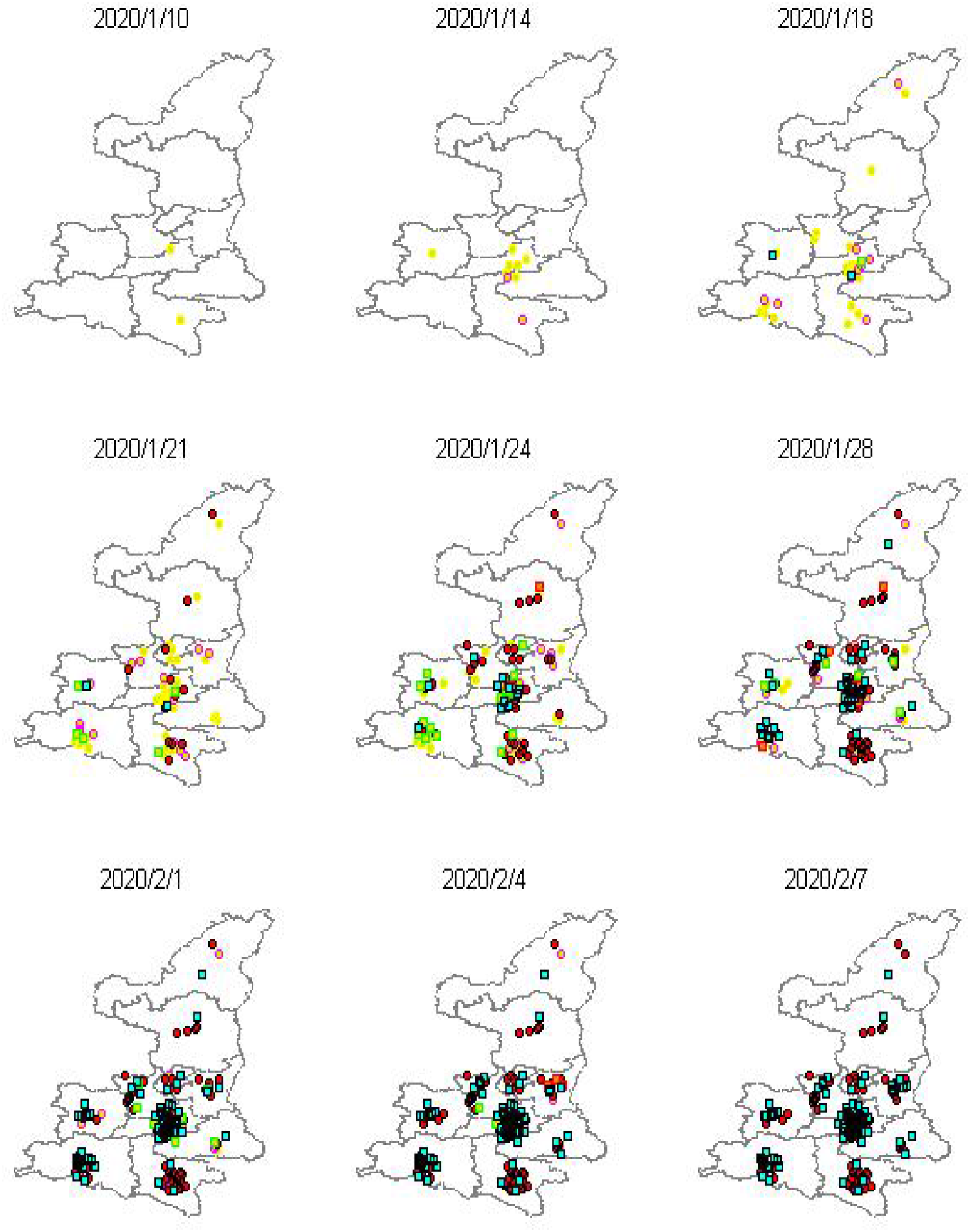
Spatial distribution of imported cases (circles) and local cases (squares) based on the traced data. The imported cases include exposed (yellow), infected (magenta), quarantined infected (red) and hospitalized (black) individuals, the local cases include infected (green), quarantined infected (red) and hospitalized (black) individuals.

The detailed records reveal how long the patients stay in latent, infected and hospitalized compartments. Based on all cases reported in Shaanxi province we use the Kaplan-Meier method to estimate the median duration from illness onset to first medical visit (quarantined, denoted by T_2_^W^) is around 0.60 days, median duration from first medical visit (quarantined) to confirmation (denoted by T_3_^W^) is around 3.43 days, shown in Figure 5. In particular, for imported cases, the median duration from importation to illness onset (denoted by T_1_^I^) is around 2.38 days with longest incubation duration of 19 days, median duration from illness onset to first medical visit (quarantined, denoted by T_2_^I^) is around 1 days with the longest (or shortest) duration of 15 (or 0) days; median duration from the first medical visit to confirmation (denoted by T_3_^I^) is around 3.05 days with the longest (or shortest) duration of 13 (or 1 days). For local cases median duration from illness onset to first medical visit (quarantined, denoted by T_2_^L^) is around 0.29 days with the longest (or shortest) infectious duration of 10 (or 0) days; median duration from the first medical visit to confirmation (denoted by T_3_^L^) is around 3.9 days with the longest (or shortest) duration of 16 (or 1 days). This analysis indicates that local people went to medical visit when having illness onset much quicker than the imported individuals. Further, these durations are relatively smaller than those for mainland China or Hubei province [?], which implies that the control measures implemented in Shaanxi are timely and medical resources are relatively sufficient.

**Figure 5.**
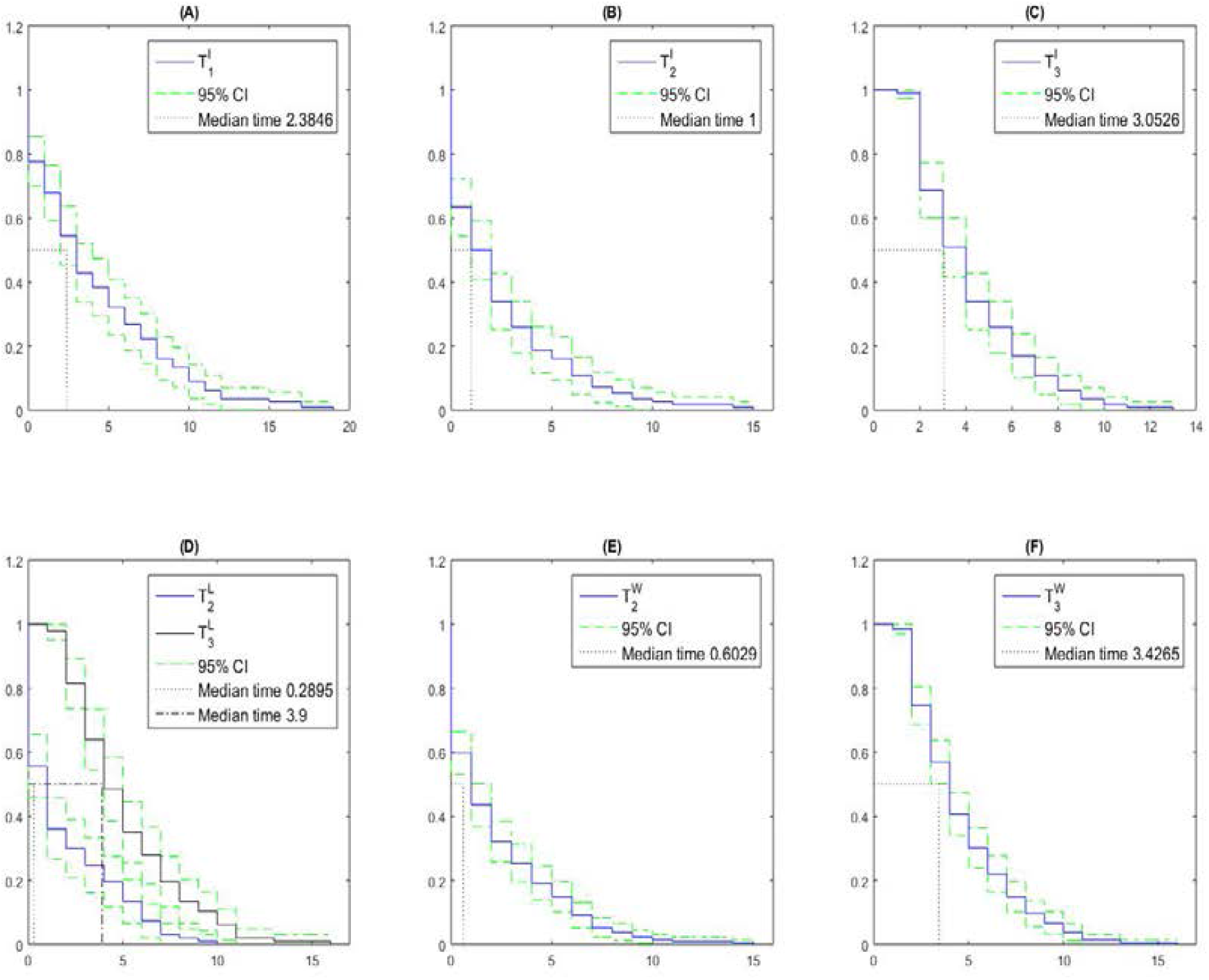
Survival analysis by using Kaplan-Meier method to estimate the duration from importation to illness onset (denoted by T_1_), the duration from illness onset to first medical visit (quarantined, denoted by T_2_) and the duration from first medical visit (quarantined) to confirmation (denoted by T_3_) for imported, local and whole cases in Shaanxi province.

### Control/ effective Reproduction numbers

From the detailed record we can find 21 transmission chains (or transmission trees) which involve 60 cases with 13 imported cases. For each transmission chain we obtained when and who infected whom, and calculated the mean control reproduction number of 1.48 with standard deviation (std) 0.98. In particular, it follows from 21 transmission chains that the number of secondary infections for each seeded case yields 2,2,1,5,1, 0,2,2,2,1, 1,1,2,1,2, 1,1,1,1,1, 1. As mentioned before, the early reported cases in Shaanxi are mainly imported cases before February 3^rd^, 2020. Then, given seed infected individuals who may get infection from January 23^rd^ to February 3^rd^, 2020, we had the transmission 13 chains (No. 1-13 as shown in Figure 6), which gives mean control reproduction number of 1.69 with std 1.18.

**Figure 6.**
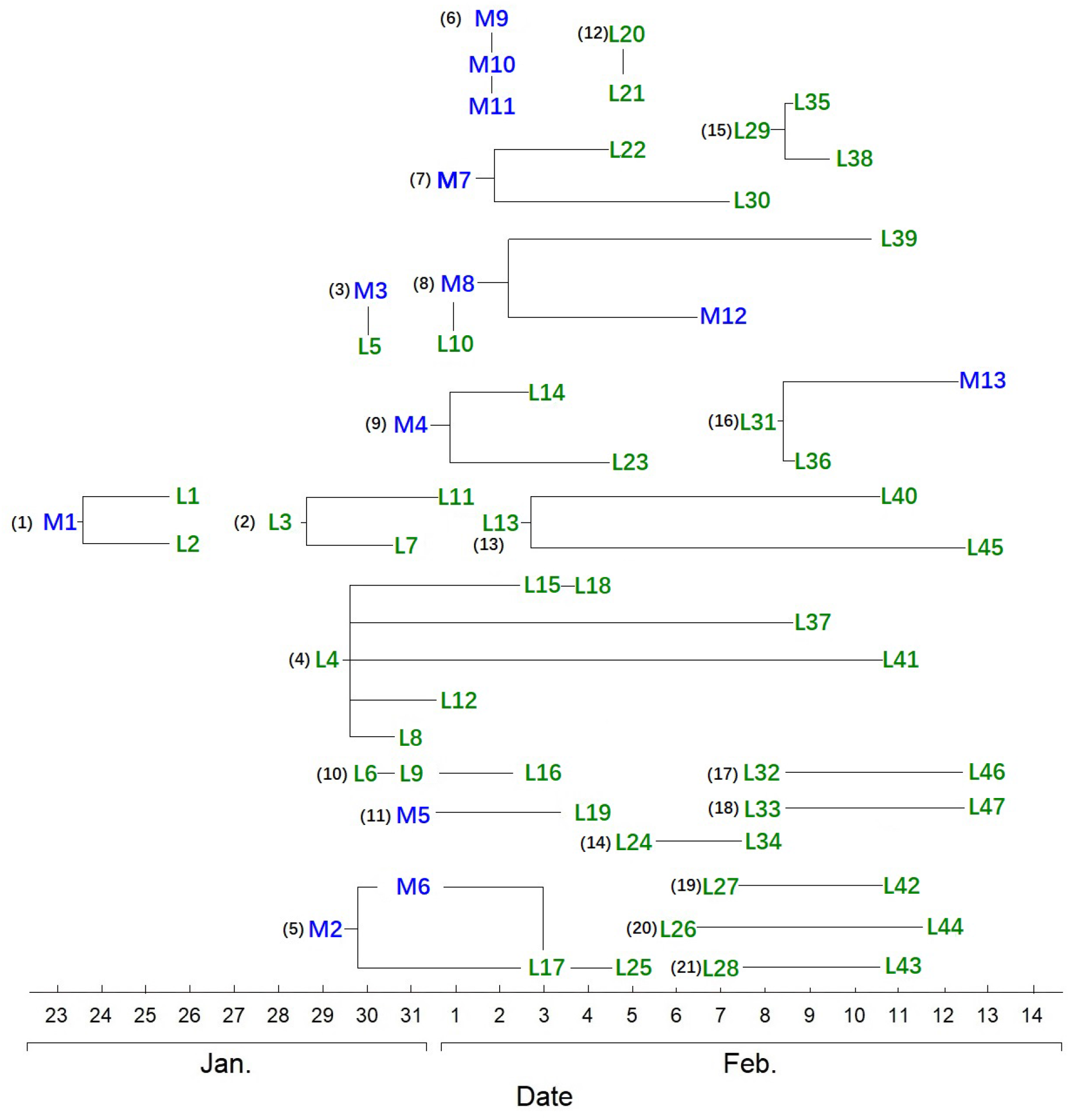
Transmission chains along the time. Blue *Mi* represent the imported cases and greens *Lj* are local cases, and *i, j* denote the identifier numbers in the system of Health Commission of Shaanxi province.

To investigate the new infections we need to know the duration from importation to confirmation (denoted by *T*_*M*_for imported cases) and the duration from infection to confirmation for local cases (denoted by *T*_*L*_). By fitting a Weibull distribution to the traced data imported cases confirmed before February 16^th^, 2020 we estimate the estimated mean and standard deviation of *T*_*M*_ as 10.68 days and 4.95 days. Let *T*_*L*_ be assumed to follow a Weibull distribution, which is the sum of the incubation period *T*_*L*1_ and the duration from illness onset to confirmation for local cases *T*_*L*2_. It follows from [3, 4, 6, 8, 11, 14, 17] that the mean and standard deviation of *T*_*L*1_ were 5.2 days and 3.91 days. The mean and standard deviation of *T*_*L*2_ were estimated to be 7.28 days and 3.89 days on the data of detailed information of local cases confirmed before February 16^th^, 2020. Then the mean and standard deviation of *T*_*L*_ was calculated to be 12.48 days and 5.52 days. Then we can estimate the daily new infections at the same period on the basis of the daily number of reported confirmed cases and the distribution of *T*_*M*_ and *T*_*L*_ by deconvolution method Figure 7(A). It is interesting to note that we can get the part of new infections before February 12^th^, 2020 from the recoded traced data in Shaanxi province, shown in Figure 7(B). It shows that the estimated new infections agree well with the traced data in terms of trend. We note that for some cases, their information is not complete, only the dates of illness onset or first medical visit are recorded, and hence they cannot be considered as new infections at certain dates. That is why the real new infections from traced data are less than the estimated ones, shown in Figure 7(B).

**Figure 7.**
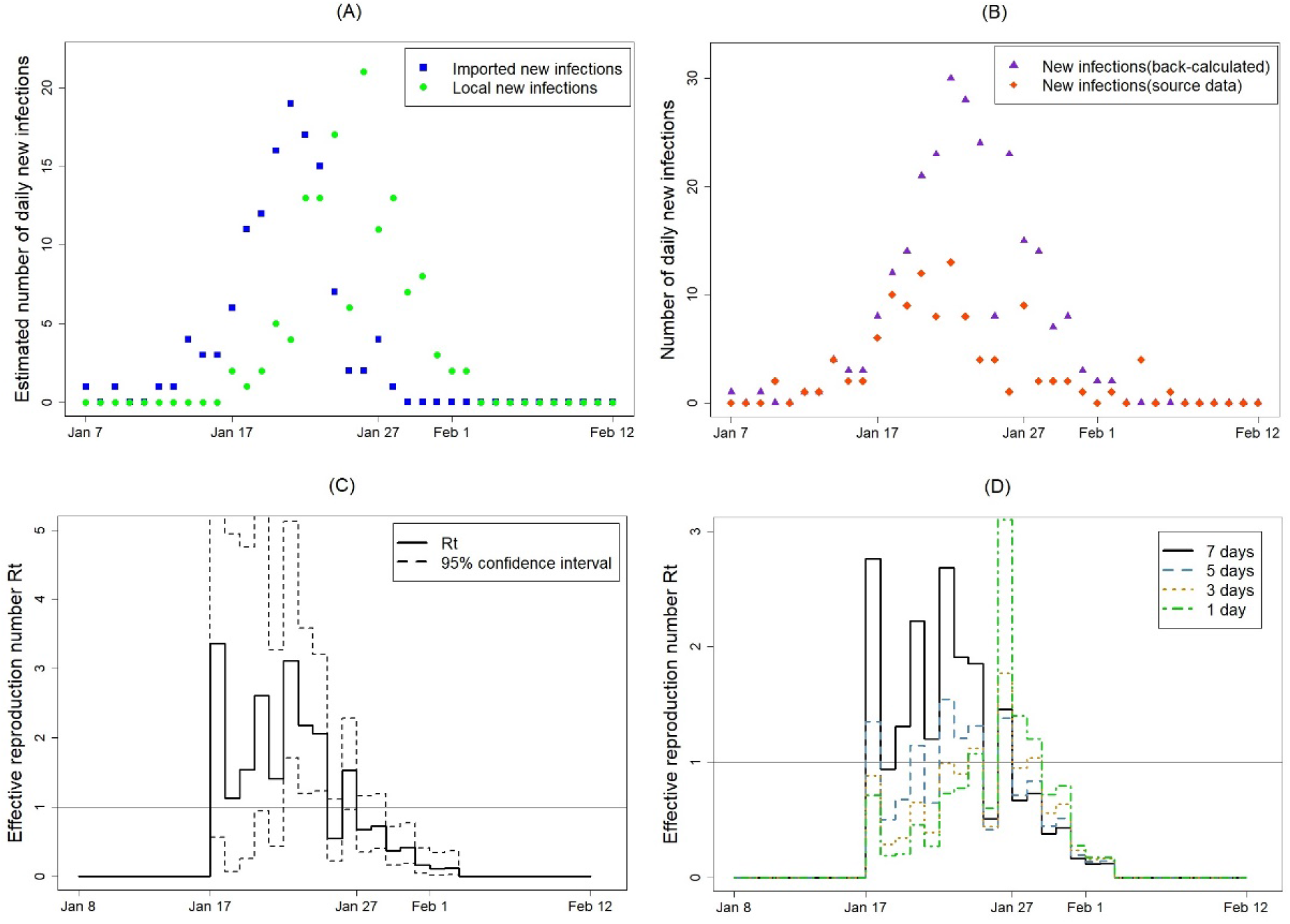
The back-calculated time series on number of new infections for imported/local individuals based on the reported confirmed cases (A), comparison of number of new infections from back calculation and traced data (B), the estimated reproduction number R(t) based on the back-calculated number of illness onset (C).

To estimate the effective reproduction number we let the distribution of serial interval *g*_*τ*_ be chosen as gamma distribution with mean of 7.5 days and standard deviation of 3.4 days [4, 11]. Because the distribution of the duration from importation of a primary imported case to infection of the secondary local case *h*_*τ*_ is unknown, we initially set *h*_*τ*_ be equal to *g*_*τ*_ and then estimate the effective reproduction number R(t), as shown in Figure 7(C). It shows the effective reproduction number started to decline on January 23^rd^, 2020 when the prediction and control strategies was strengthened, was less than 1 since around January 27^th^, 2020, and stabilized at almost 0 recently, which means few new infections occur. In order to investigate the variation of the effective reproduction number with *T*_*h*_ varies, sensitivity analysis was carried out by changing the mean of *T*_*h*_ from 1 to 7 days, as shown in Figure 7(D). It shows that the more the mean of *T*_*h*_, the greater duration of the R(t) being greater than 1 and the earlier the R(t) exceeds the unity.

### Does resumption of work induce the second outbreak

Since the Spring Festival holiday most people have been at home, when do they return back to work/study is a challenging problem. Consequently, we investigate the effect of population movement on infections, especially on the possibility of inducing the second outbreak. To this end, we initially estimate the unknown parameters by least square methods, listed in Table 1, by fitting the discrete stochastic model to the traced data between January 10^th^ to February 7^th^ (purple curve with dots), and by using the mean parameter values of 1000 times of least square estimations (listed in Table 1) we get 500 stochastic trajectories of model (1), as shown in Figure 8(A-C). Figure 8(D) shows the estimated susceptible individuals vary significantly. Mean estimated values (green curves) and 95% confidence interval (CI) of *E*_*t*_, *I*_*t*_, *H*_*t*_ compartments are shown in Figure 8(E-G). Note that on February 4^th^, 2020 the first cured individual was covered, and the estimated recovery rate *γ*_*H*_ was so small that mean value of *H*_*t*_ persistently and slowly increases, shown in Figure 8(C). However, tripling the recovery rate can induce the value of *H*_*t*_ decline. By using the data between January 10^th^ to February 16^th^, 2020 (purple curve with dots) we verify the model and find the traced data are within the 95% CI of estimated *E*_*t*_, *I*_*t*_ and *H*_*t*_. It is worth mentioning that the number of suspected cases *B*_*t*_ (upper green curve in Figure 8(D)) quickly increases since people, once having illness-like symptom or fever, were likely access to medical visit and were considered to be suspected individuals and isolated. While the case-driven cumulative quarantined individuals via contact tracing (lower green curve in Figure 8(D)) becomes to stabilize, and is less than the number of suspected individuals due to relatively small number of confirmed cases. The total number of these two classes indicates the prevention and control strategies implemented in Shaanxi province are strong.

**Figure 8.**
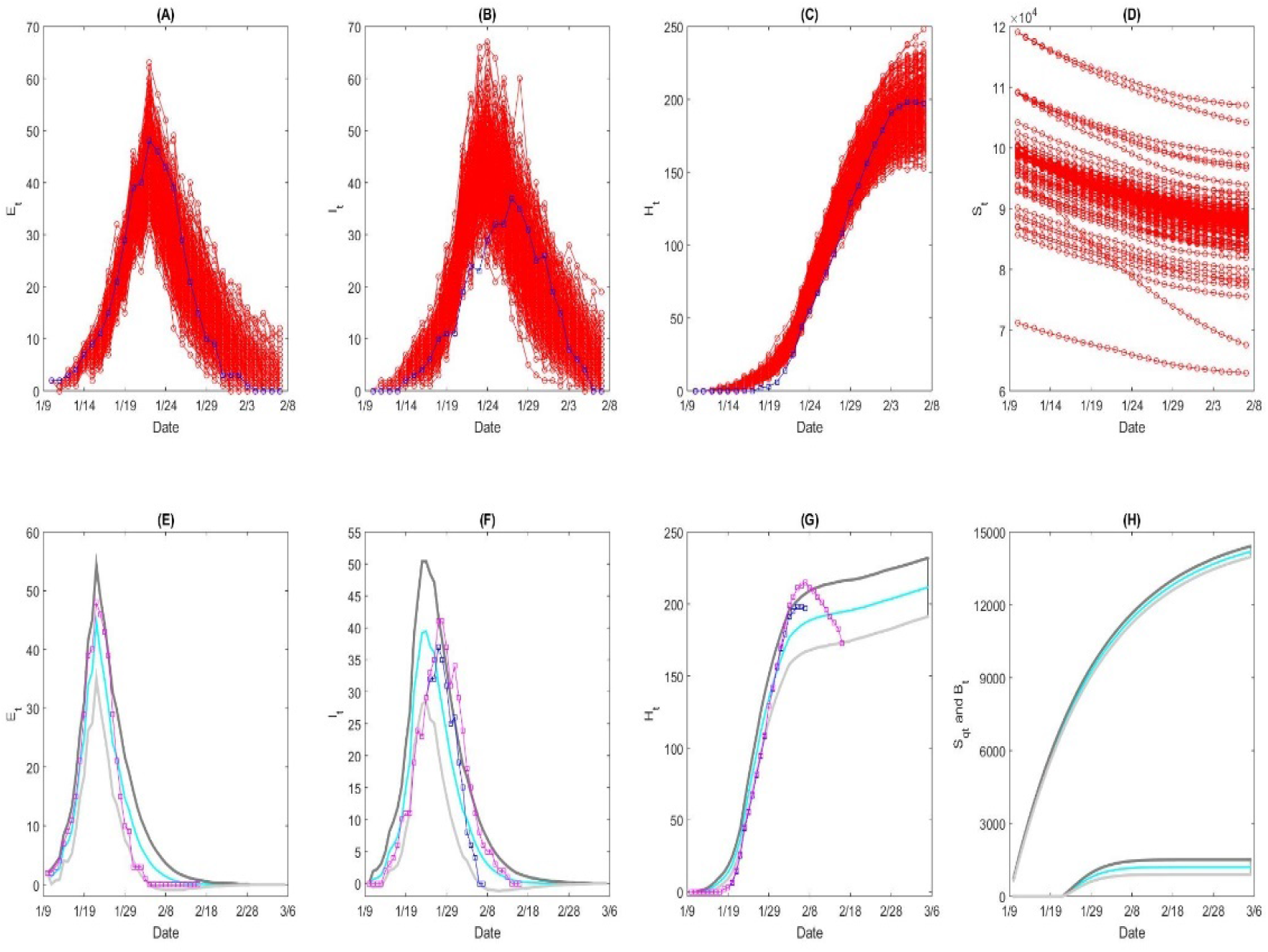
Data fittings with 500 simulations (A-C). The variations of the number of susceptible population are significantly (D). The mean values (green curve) and 95% CI (grey curve) for E(t), I(t) and H(t) compartments (E-G). D: the number of suspected individuals B(t) (upper green curve) and the cumulative number of the quarantined individuals (lower green curve).

To investigate the effect of resumption of work on the epidemic, especially on possible second outbreak, by which we may suggest the public health sectors when workers can return back to Shaanxi, we predicate the disease infection by using the data on imported cases and moving from other cities to Shaanxi province (shown in Figure 2). Suppose population to start to return back to Shannxi province from February 24^th^ to March 9^th^, 2020, we can see from Figure 9 (A-B) that there is a large second outbreak. If the proportions of imported cases are decreased by 30%, 60% and 90%, the second outbreak becomes smaller and smaller, shown in Figure 9(C-D), (E-F), and (G-H), respectively. Further, if the inflow mode is changed into the intermittent inflow, that is, population are allowed to move into Shaanxi every other week, we can from Figure 10 that there are one more outbreaks with low peak values. Decreasing the imported proportions greatly reduces the peak values of the following outbreaks. Comparing Figure 9 and Figure 10 indicates that intermittent inflow mode may induce small outbreaks and a relatively good choice for resumption of work.

**Figure 9:**
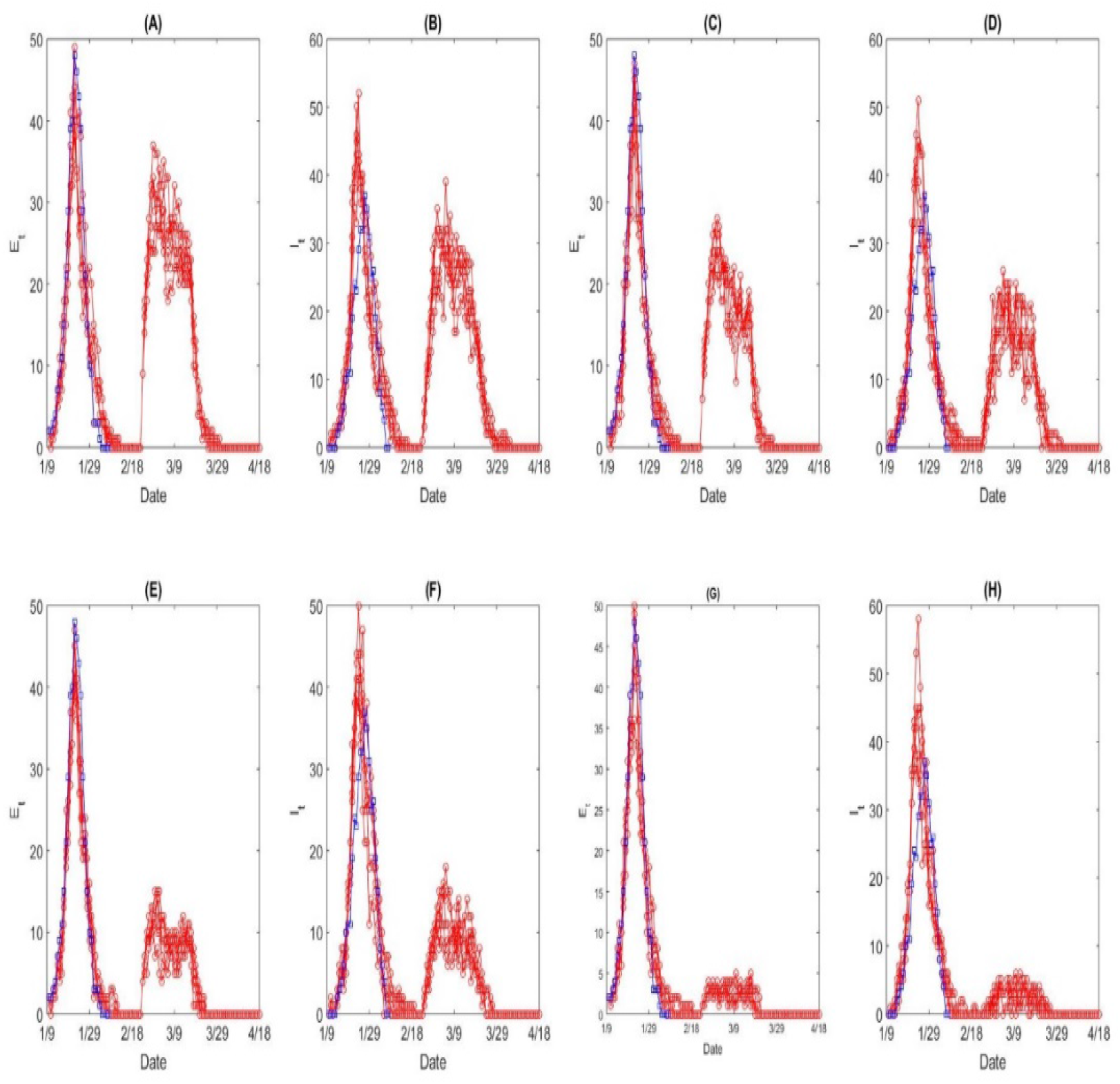
Effect of continuous inflow on the number of exposed individuals (A) and infected individuals (B) with baseline parameter values, with 30% decreased inflow (C-D), with 60% decreased inflow (E-F), with 90% decreased inflow (G-H). Here the inflow has been lasting for 24 days (from February 24^th^ to March 18^th^).

**Figure 10:**
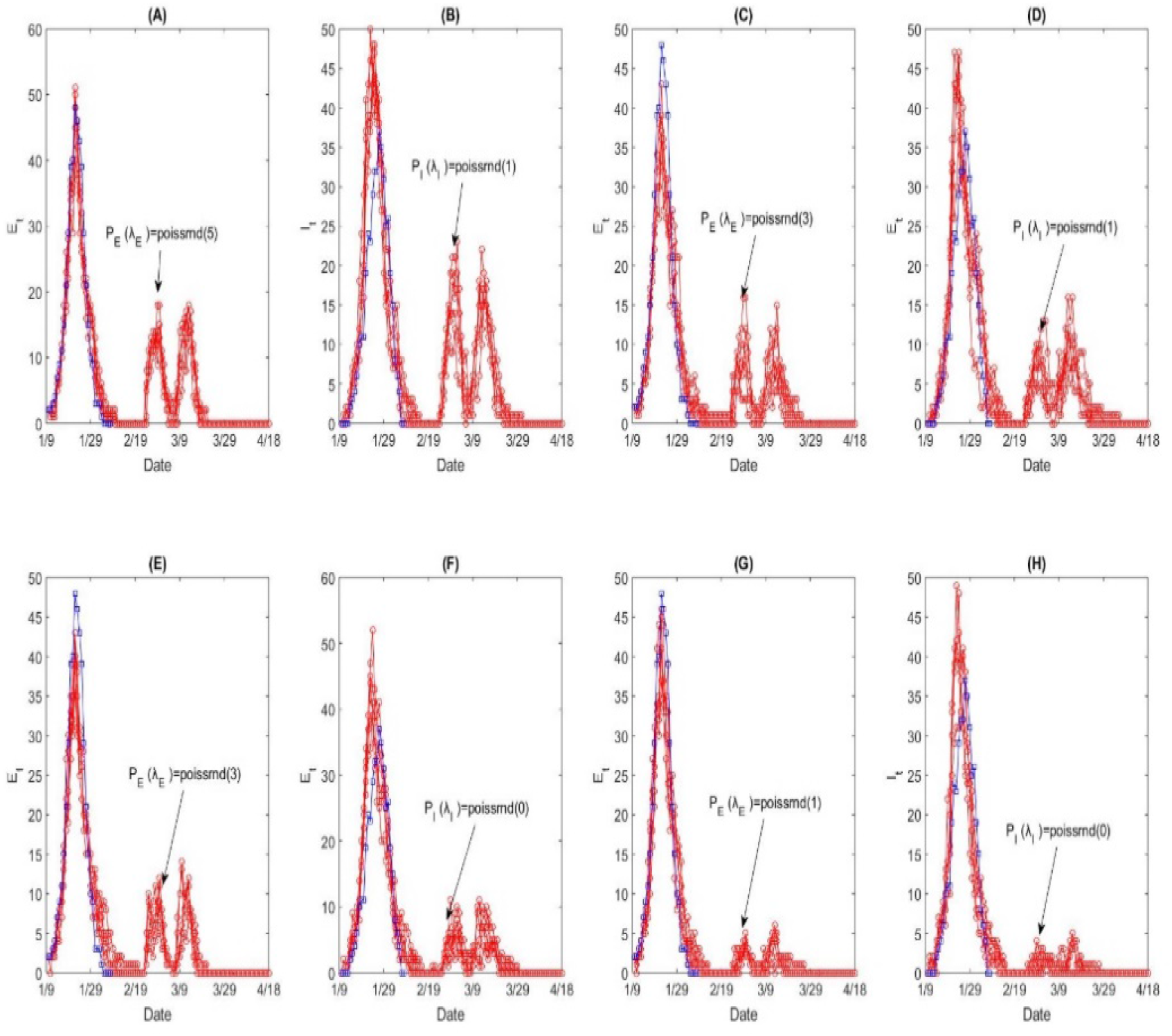
Effect of intermittent inflow on the number of exposed individuals (A) and infected individuals (B) with baseline parameter values, with 30% decreased inflow (C-D), with 60% decreased inflow (E-F), with 90% decreased inflow (G-H). Here the inflow is on for the first (from February 24^th^ to March 1^st^) and third weeks (from March 9^th^ to 15^th^) and off for the second and fourth week and the following days.

## Discussion

Shaanxi province is located northwest of China, adjacent to Hubei in the south. Therefore, except for Xi’an city, the capital of Shaanxi, many confirmed cases are in the southern part of Shaanxi province. It is interesting to note that Health Commission of Shaanxi Province released the case number every day with very detailed information, by which we can know when and who infected whom. Then except for the new reported (or cumulative) number of confirmed cases, we can get the daily number of individuals who are in latent (or quarantined latent), infected and hospitalized compartments, which gives the evolution process of the epidemic in Shaanxi province. We can get the durations from illness onset to the first medical visit, to the hospitalization, and to confirmation of the case. These durations are relatively smaller than those for mainland China or Hubei province [3, 4, 6, 8, 11, 14, 17], which implies that the control measures implemented in Shaanxi are timely and medical resources are relatively sufficient.

From the detailed data description, we can obtain the transmission chains over the time, which enable to calculate the control reproduction number under the implementation of stringent control strategies. The mean control reproduction number ranges from 1.48 to 1.69, and the distribution of these chains agrees well with the effective reproduction number. Note that the effective number has been declining, and is less than unity since January 27^th^ 2020. Recently it has stabilized at almost zero, meaning that no new infections occur, which seem to be in line with real data, as shown in Figures 1 and 7 [7].

In this study we extend our deterministic model framework [9, 18-20] to a discrete system incorporating stochastic modeling of imported cases to represent the relatively small number of cases observed in the Shaanxi province, with about half of them being imported cases. It is worth noting that traced data is around 7 days ahead of the reported ones, as illustrated in Figure 1(A). Moreover, detailed data records revealed that the time interval between the first visit to a doctor and confirmation was longer, with a median of around 4 days. Then, the number of reported cases is not suitable to identify the model or estimate the parameters due to delays such as report delay, confirmation delay, etc. Therefore, it is not accurate to associate the number of confirmed cases with the number of cases in hospital, which may seriously overestimate the scale of the epidemic. Therefore, the multiple traced data sets have been employed to identify the proposed discrete stochastic model and estimate the unknown parameters in this work. Consequently, we use the number of individuals in E, I, and H classes form traced data rather than the newly (or cumulative) reported number to parameterize the model.

We investigate the impact of population movement (resumption of work) on the potential second outbreak by using movement data from other cities to Shaanxi province. Our main results indicate that the proportion of imported cases is a key variable in order to determine whether the second outbreak is likely or not to occur. The greater the proportion of imported cases the larger the second outbreak has. Furthermore, intermittent importation may induce one further outbreak episode but with small peak values.

From our study, we can conclude that it is very important to study the epidemic situation of COVID-19 by using data from contact tracing rather than simply using the number of confirmed cases, especially in the areas where the imported cases are the main source of infection. This can not only provide important data information for the accurate evaluation of the epidemic situation in other areas outside Hubei Province, but also provide important and accurate data supporting the evaluation of the effectiveness and timeliness of public health intervention strategies in different areas.

In the present investigation, we devised a stochastic model that can be used as an important model framework and methodology for the evaluation of imported cases on local epidemic in the Shaanxi province or other similar provinces. From the data analysis, it seems that the stringent public health interventions adopted by the Chinese authorities are effective in controlling the infection. According to data from contact tracing, the first imported cases in the Shaanxi Province occurred earlier than January 10^th^ 2020, and most of the imported cases were imported to the Shaanxi Province before the implementation of the national travel restrictions on January 23^rd^ 2020. Therefore, the earlier the implementation of the travel restrictions, the greater the reduction of infected cases by COVID-19 in the Shaanxi province has. An intermittent population inflow with low proportion of imported cases may significantly mitigate the transmission risk. Thus, if the public health authorities can strictly monitor and reduce the number of imported cases, it would be possible for the Shaanxi province to return back and resume working continuously.

## Conclusions

It is very important to study the epidemic situation of COVID-19 by using traced data rather than simply using the number of confirmed cases, especially in the areas where the imported cases are the main and the epidemic situation is not serious. This can not only provide important data information for the accurate evaluation of the epidemic situation in other areas outside Hubei Province, but also provide important and accurate data support for the evaluation of the effectiveness and timeliness of public health intervention strategies in different areas.

We proposed a stochastic model which can provide an important model framework and methodology for the evaluation of imported cases on local epidemic in the Shaanxi province (and can eventually be adapted for other similar provinces of China). Data analysis reveals that strengthening the public health interventions, tracing imported cases and improving the confirmation rate are effective and timely after January 23^rd^ 2020. According to the traced data shown in Figure 1, the first imported cases in Shaanxi Province were earlier than January 10, and most of the imported cases were imported to Shaanxi Province before the national travel restrictions were implemented on January 23nd, as shown in Figure 2. Therefore, the earlier implementation of travel restriction will greatly reduce the epidemic situation of COVID-19 in Shaanxi province, which was not serious.

The findings suggest that keeping an intermittent population inflow with a low proportion of imported cases may induce a small second outbreak and hence decrease the transmission risk. Thus, if we can strictly monitor and reduce the number of imported cases, it is possible for Shaanxi to resume work continuously, especially intermittently.

## Data Availability

Health Commission of Shaanxi Province. Available at http://sxwjw.shaanxi.gov.cn/col/col863/index.html

## Acknowledgement

This research was funded by the National Natural Science Foundation of China (grant numbers: 11631012 (YX, ST), 61772017 (ST)), and by the Canada Research Chair Program (grant number: 230720 (JW) and the Natural Sciences and Engineering Research Council of Canada (Grant number:105588-2011 (JW).

## Notes

### Competing Interest Statement

The authors have declared no competing interest.

